# Integrative Bioinformatics Analysis Identifies Noninvasive miRNA Biomarkers for Lung Cancer

**DOI:** 10.1101/2020.12.31.20248963

**Authors:** Andrew Gao

## Abstract

Non-small cell lung cancer (NSCLC), a subtype of lung cancer, affects millions of people. While chemotherapy and other treatments have improved, the 5 year survival rate of NSCLC patients is still only 21%. Early diagnosis is essential for increasing survival as treatments have higher effectiveness at earlier stages of NSCLC. Noninvasive blood-based liquid biopsy tests for NSCLC may be useful for diagnosis and prognosis. MicroRNA (miRNA) and messenger RNA present in blood can serve as biomarkers for such tests. The present study identified 13 miRNAs that are underexpressed in the tissue and blood of NSCLC patients using Gene Expression Omnibus data. Following Kaplan Meier analysis, miR-140-3p, miR-29c, and miR-199a were selected as candidate biomarkers and demonstrated statistically significant prognostic power. An ROC analysis of miR-140-3p expression between NSCLC patients and controls had an area under curve value of 0.85. Functional enrichment analysis of the miRNA target genes revealed several overrepresented pathways relevant to cancer. Eight target genes were hub genes of the protein protein interaction network and possessed significant prognostic power. A combination of IL6, SNAI1, and CDK6 achieved a hazard ratio of 1.4 with p < 0.001. These biomarkers are especially valuable because they can be identified in blood and reflect the tumor state. Since all miRNAs were underexpressed in both tissue and blood, detecting expression of a biomarker miRNA in blood provides information on its expression in tissue as well. These miRNAs may be useful biomarkers for NSCLC prognostic and diagnostic tests and should be further studied.

## Introduction

Lung cancer, the leading cause of cancer mortality, is responsible for millions of deaths globally. In 2018, an estimated 1.8 million people died from lung cancer and approximately 2.1 million new cases were diagnosed worldwide [1]. The primary subtype of lung cancer is non-small cell lung cancer (NSCLC), which comprises more than 80% of all lung cancer cases [2]. NSCLC consists mainly of two subgroups: lung adenocarcinoma (LUAD) and squamous cell carcinoma (LUSC). While treatments for NSCLC have improved, the mortality rate is still high: the 5-year survival rate of patients with NSCLC is only 21% [3]. This is often attributed to a lack of early diagnosis. Currently, NSCLC is generally diagnosed at late stages when the cancer has already progressed significantly and patient prognosis is poor. At advanced stages, NSCLC is much more difficult to treat. In order to improve NSCLC survival rates, it is imperative to diagnose NSCLC at early stages.

Identification of diagnostic and prognostic biomarkers for NSCLC is necessary for the development of accurate tests. Biomarkers are present in a variety of mediums, from cerebrospinal fluid to tissue to saliva. Blood and its components, including plasma and serum, are attractive sources of cancer biomarkers, because of the non-invasive nature of blood collection. Several types of biologically relevant biomarkers are present in blood, such as metabolites, circulating tumor DNA, and microRNAs (miRNA) [4].

MiRNAs are a type of non-coding RNA (ncRNA) that are involved in the regulation of gene expression. They function by binding to target messenger RNAs (mRNA) which silences expression. Generally 20-25 nucleotides in length, miRNAs have been implicated in cancer as they are relevant in apoptosis, cell proliferation, and other tumor-associated processes [5-7]. Previously, several differentially expressed miRNAs (DEMiRNAs) have been identified in NSCLC blood and tissue [8-10]. DEMiRNAs in blood have potential for serving as biomarkers in noninvasive diagnostic and prognostic tests.

Blood-based miRNAs can be profiled using various methods, including RNA-seq and microarray profiling. Previously, microarray-based miRNA profiling has been used to select biomarkers for other diseases, such as breast cancer [11]. Several studies have identified differentially expressed miRNAs in NSCLC blood that could serve as biomarkers. For example, Xue et al. reported that miR-1228-3p and miR-181a-5p were potential biomarkers for lung cancer in serum samples [12].

While miRNA expression profiling is highly useful, there are often significant discrepancies in DEMiRNAs between studies as a result of different methodologies, sample characteristics, age of samples, and other factors. MiRNAs found to be significantly differentially expressed by one study are often not found in another. Results can be highly variable due to factors such as different profiling platforms or procedural differences [13]. Having a reliable panel of miRNAs is imperative, especially when dealing with NSCLC, a heterogeneous disease.

In the present study, three blood miRNA profiling datasets and one tissue miRNA profiling dataset were analyzed to identify differentially expressed miRNAs (DEMiRNAs) consistently dysregulated in blood and tissue. Target gene prediction was performed on overlapping miRNAs and the resulting genes were analyzed in Gene Ontology (GO) and the Kyoto Encyclopedia of Genes and Genomes (KEGG). A protein-protein interaction (PPI) network was constructed and analyzed to elucidate the molecular mechanisms of the targeted genes and their role in NSCLC carcinogenesis. Survival analysis was conducted on hub genes. Finally, survival analysis, receiver operating characteristic (ROC) analysis, and methylation analysis were performed on candidate biomarker miRNAs to validate their relevance in NSCLC.

## Methods

### Dataset Selection

The Gene Expression Omnibus (GEO) is a publicly available online database that contains gene expression data from various studies [14-15]. GEO was searched for datasets matching {“blood” or “serum” or “plasma”}, “NSCLC” (and word forms), “*Homo Sapiens”*, and “miRNA profiling”. Studies GSE137140, GSE93300, and GSE94536 were selected for analysis [16-18]. GSE137140 contained serum miRNA profiling data while GSE93300 and GSE94536 contained plasma miRNA profiling data. Another dataset, GSE53882, containing tissue miRNA data from paired samples, was selected as well [19]. The same selection criteria were applied except for “tissue” instead of {“blood” or “serum” or “plasma”}. All four studies used microarrays to collect data. Microarrays utilize oligonucleotide probes to bind and capture complementary strands of DNA (cDNA). The cDNA is created by reverse transcribing the miRNA isolated from the sample.

**Table 1:** Information on GSE137140, GSE93300, GSE94536, and GSE53882, including number of NSCLC and control samples and sample source.

| Dataset ID | Number of NSCLC samples | Number of control samples | Source | Profiling Type | Platform |
| --- | --- | --- | --- | --- | --- |
| GSE137140 | 1566 | 1774 | Serum | miRNA | GPL21263 |
| GSE93300 | 9 | 4 | Plasma | miRNA | GPL21576 |
| GSE94536 | 6 | 3 | Plasma | miRNA | GPL21576 |
| GSE53882 | 397 | 151 | Tissue | miRNA | GPL18130 |
| Total | 1978 | 1932 | NA | NA | NA |

### Differential Expression Analysis

The five datasets were screened for DEMiRNAs in RStudio using the programming language R and the Limma package [20]. P values (t test) and fold change was calculated for each miRNA based on data from the studies’ Series Matrix Files. The online tool GEO2R, provided by the Gene Expression Omnibus, was used to verify a normal distribution of gene expression values. Quantile normalization was applied if necessary. The statistical significance cutoff for differential expression was set as p < 0.05. The DEMiRNAs were separated based on overexpression or underexpression (logFC>0 or logFC<0). LogFC is the log 2 variant of FC, or fold change. MiRNAs that were statistically significantly differentially expressed in all four datasets were identified and sorted by magnitude of fold change. Venn diagrams of the DEMiRNAs in each dataset were constructed via Jvenn software [21]. LogFC data from the four datasets was retrieved for each DEMiRNA using Python and a heatmap was constructed using Graphpad Prism.

### Target Gene Prediction and Functional Enrichment Analysis

Target genes of the DEMiRNAs were predicted using three online databases in order to increase prediction accuracy. MiRWalk, miRDB, and mirTarBase were utilized to predict target genes [22]. Only target genes identified by all three databases were kept for further analysis. The target genes were analyzed via Gene Ontology (GO) [23]. GO is a large database that stores information on biological pathways, components, and functions, and the involved genes. GO’s tool PANTHER processes a provided list of genes and calculates which biological entities those genes are overrepresented in [24]. The list of target genes was submitted to PANTHER and enriched biological processes and molecular functions were received. The Fisher’s Exact statistical test was used to determine statistical significance (p<0.05) with the false discovery rate (FDR) used for correction. P < 0.05 was regarded as the significance cutoff for overrepresentation.

### Network Analysis

The list of target genes was analyzed in the Search Tool for the Retrieval of Interacting Proteins (STRING) to construct a network of the protein-protein interactions of the gene products [25]. STRING is a bioinformatics tool that scans several protein databases to create a network visualization of input genes and how their protein products interact. Additionally, STRING calculates statistically significant Gene Ontology processes and pathways, similar to PANTHER. The confidence level of protein-protein interactions was set to highest confidence (0.9) and solitary node genes were hidden to improve graph readability. A tab separated value file (TSV) containing the entire unfiltered graph was downloaded and imported into Cytoscape [26]. Cytoscape is a desktop software tool that can perform powerful statistical analyses on networks. The Network Analyzer tool in Cytoscape was used to calculate metrics of the gene network such as node degree and clustering coefficient [27]. Genes with a degree of greater than or equal to 20 were considered hub genes. The degree is the number of genes connected to a certain gene. Additionally, the Molecular Complex Detection MCODE plugin was used within Cytoscape to discover significant modules in the protein interaction network. MCODE is a graph clustering algorithm that is able to identify “densely connected regions” in networks [28].MCODE was set to the following settings: degree cutoff = 2, node score cutoff = 0.2, K-core = 2, and depth = 100. The STRING plugin was used to perform functional enrichment analysis on individual modules.

### Analysis of Hub Genes

The Kaplan-Meier plotter tool was utilized to calculate and visualize NSCLC survival curves for lung adenocarcinoma and squamous cell carcinoma patients [29]. The Kaplan-Meier plotter tool combines gene expression data from The Genome Cancer Atlas (TCGA), the Gene Expression Omnibus, and the European Genome-Phenome Archive. The hub genes identified in Cytoscape were submitted to the Kaplan-Meier tool and the survival curves were studied. The settings were: median split of patients and overall survival. Logrank P values and hazard ratios using 95% confidence intervals were calculated for each gene. Promising hub genes were analyzed using the GEPIA tool [30].

### MiRNA Biomarker Selection

Kaplan-Meier survival curves were generated for the differentially expressed miRNAs with the settings as median split of patients and overall survival metrics. MiRNAs with statistically significant prognostic power according to the log rank p value were further analyzed in MiRTV, a tool that visualizes expression of miRNAs based on TCGA datasets [31]. MiRTV was used to validate differential expression in cancer tissue for each miRNA. Plots of miRNA expression in LUAD and LUSC were generated and the log2 transformation was applied. TCGA Wanderer software was applied to analyze differential methylation of candidate miRNAs to evaluate the influence of methylation in NSCLC gene expression [32]. GraphPad Prism was used to perform ROC analysis.

## Results

### Differentially Expressed miRNA Identification

GSE53882, GSE137140, GSE93300, and GSE94536 were selected for analysis. Gene expression value distribution was normal in all datasets except GSE93300. Quantile normalization was performed for dataset GSE93300. No statistically significant overexpressed miRNAs were identified by GSE94536.

**Table 2:** Number of overexpressed and underexpressed DEMiRNAs in each dataset.

| Dataset ID | Number of overexpressed miRNAs (logFC>0) | Number of underexpressed miRNAs (logFC<0) |
| --- | --- | --- |
| GSE137140 | 280 | 2200 |
| GSE93300 | 2161 | 58 |
| GSE94536 | 0 | 242 |
| GSE53882 | 550 | 564 |

**Figure 1:**
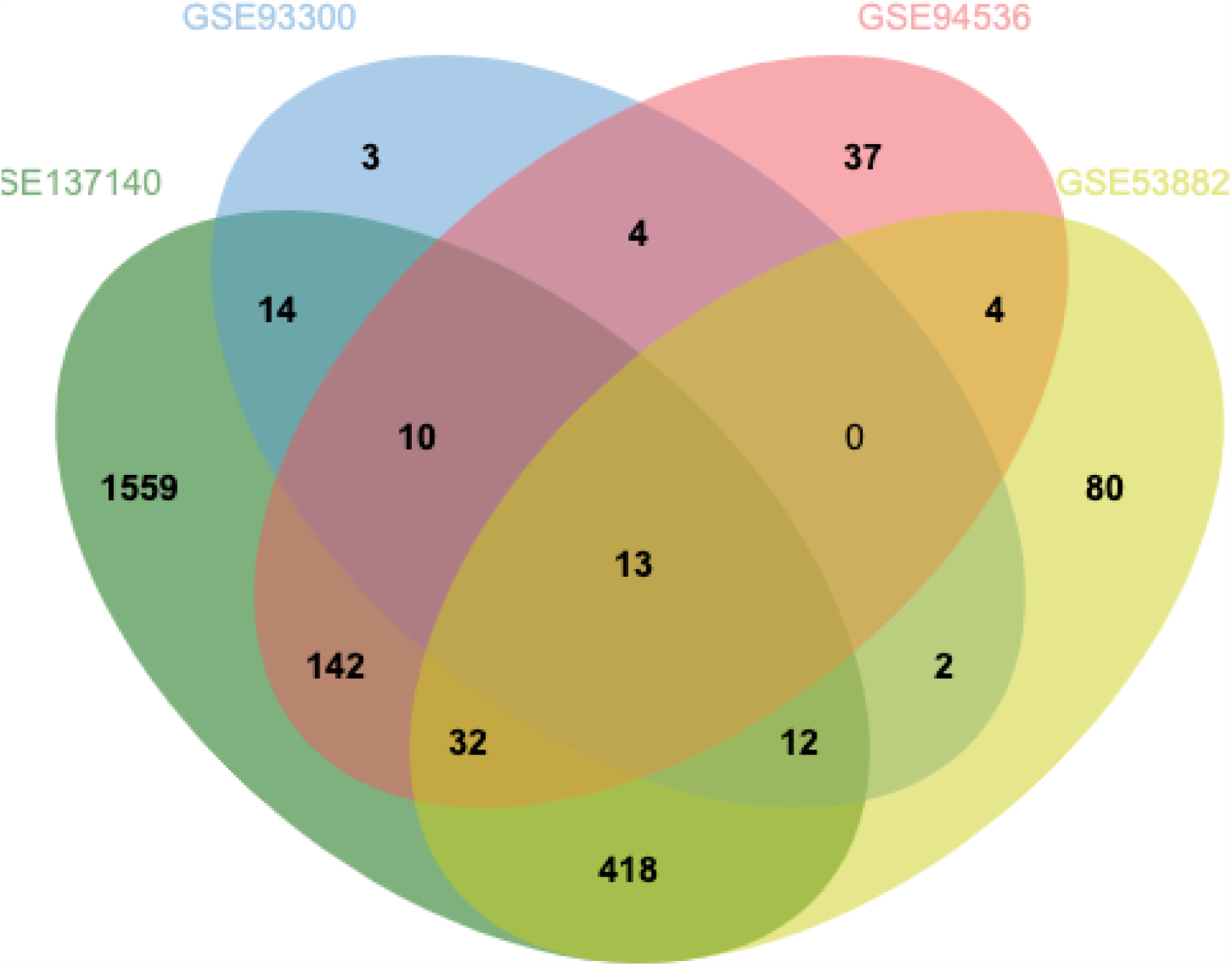
Four way Venn diagram displaying overlapping DEMiRNAs between GSE137140, GSE93300, GSE94536, and GSE53882. Thirteen DEMiRNAs were common across all four datasets.

The underexpressed DEMiRNAs from each of the four datasets were inputted into the tool JVenn to generate a four-way Venn diagram and identify overlaps. Thirteen DEMiRNAs were present in all four datasets and fifty four DEMiRNAs were found in three out of four datasets. The majority of DEMiRNAs (1679 out of 2330 unique DEMiRNAs) were only present in one out of four datasets, despite the fact that all DEMiRNAs had p values of less than 0.05. The thirteen overlapping DEMiRNAs were selected for further analysis and their logFC values were retrieved from each dataset.

**Figure 2:**
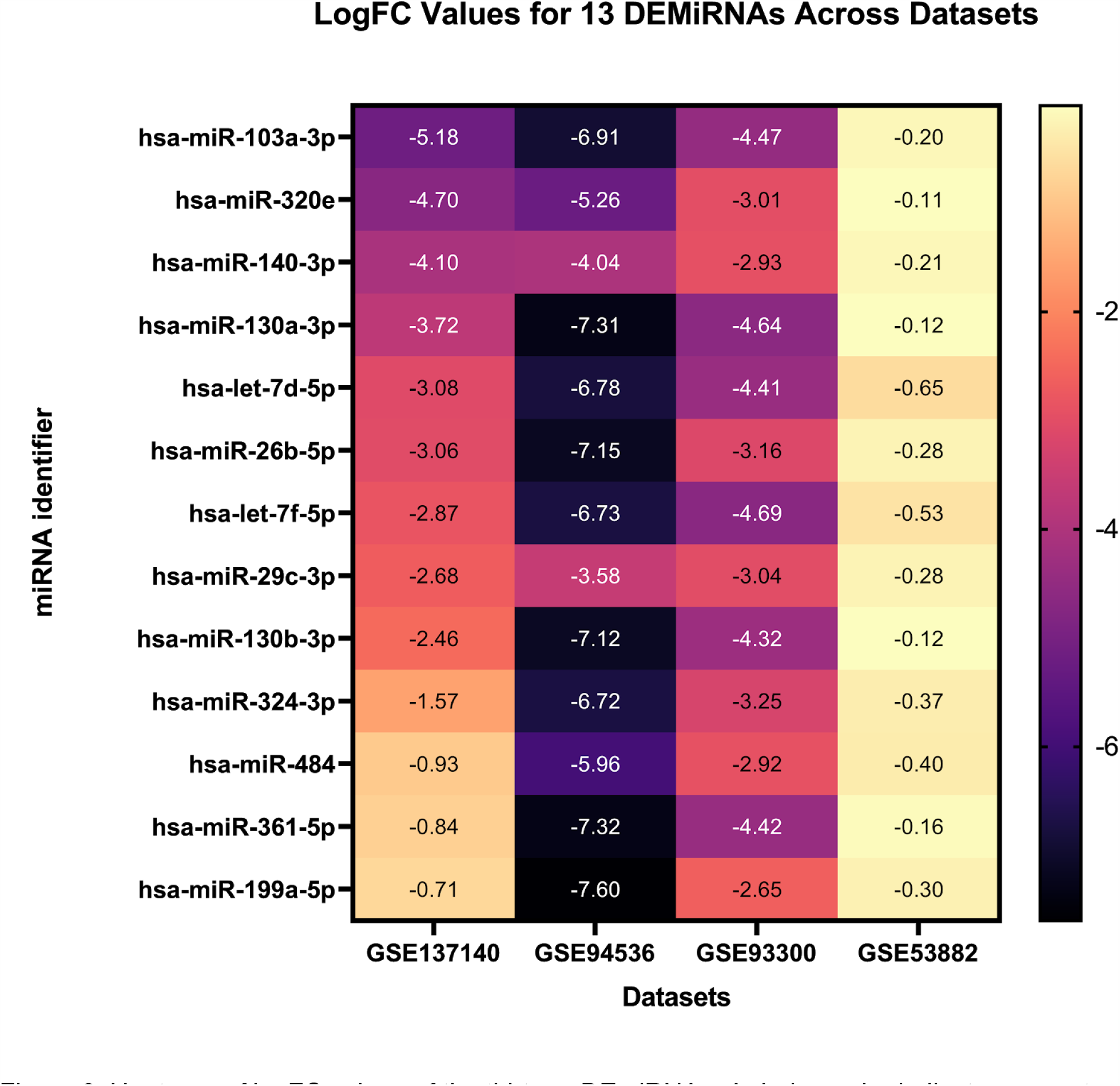
Heatmap of logFC values of the thirteen DEmiRNAs. A darker color indicates a greater magnitude of downregulation (more negative). The DEmiRNAs are plotted in order of greatest negative value based on their logFC in GSE137140. GSE53882 had notably less extreme logFC values than the other datasets, however this may be attributed to experimental differences. All logFC values displayed on the heatmap are statistically significant (p<0.05).

### Target Gene Prediction and Functional Enrichment Analysis

In total, 349 target genes were identified for the 13 DEMiRNAs through the unanimous consensus of miRwalk, mirTarBase, and miRdb. These genes are theoretically overexpressed in NSCLC because the miRNAs that regulate them are underexpressed. PANTHER analysis of the target genes revealed several significantly enriched Gene Ontology biological processes and molecular functions. The top fifteen overrepresented biological processes and functions are displayed. Several are commonly implicated in cancer development, proliferation, invasion, and metastasis,

The most enriched biological process was “positive regulation of epithelial to mesenchymal transition involved in endocardial cushion formation” (GO:1905007). Several other biological processes related to the epithelial to mesenchymal transition were also significantly enriched. The process of “pathway-restricted SMAD protein phosphorylation” was enriched by 18.23 times. Meanwhile, the process of “heterochromatin assembly” was enriched by 13.16 times. Notably, the process of “negative regulation of gene silencing by miRNA” was enriched by 12.47 times. In general, overrepresented biological functions were related to the epithelial to mesenchymal transition, gene silencing by miRNAs, response to cholesterol, heterochromatin, and cardiac cell growth and proliferation.

Several overrepresented molecular functions were also identified. These molecular functions were generally related to protein kinase activity, transcription regulatory activity, chromatin binding, and transcription factor binding. The most enriched molecular function was GO:0051575: 5’-deoxyribose-5-phosphate lyase activity.Finally, a PANTHER analysis of Gene Ontology cellular components showed that the RISC complex was enriched by 16.92 times.

**Table 3:** Overrepresented Gene Ontology biological processes ranked by fold enrichment value.

| GO biological process complete | Number of Genes in List | Expected Number of Genes | Fold Enrichment | P Value | FDR |
| --- | --- | --- | --- | --- | --- |
| positive regulation of epithelial to mesenchymal transition involved in endocardial cushion formation (GO:1905007) | 3 | 0.08 | 35.54 | 2.39E-04 | 1.68E-02 |
| regulation of epithelial to mesenchymal transition involved in endocardial cushion formation (GO:1905005) | 3 | 0.1 | 29.62 | 3.54E-04 | 2.24E-02 |
| positive regulation of cardiac epithelial to mesenchymal transition (GO:0062043) | 3 | 0.14 | 22.21 | 6.78E-04 | 3.60E-02 |
| regulation of cardiac epithelial to mesenchymal transition (GO:0062042) | 3 | 0.15 | 19.75 | 8.93E-04 | 4.36E-02 |
| pathway-restricted SMAD protein phosphorylation (GO:0060389) | 4 | 0.22 | 18.23 | 1.50E-04 | 1.16E-02 |
| heterochromatin assembly (GO:0031507) | 6 | 0.46 | 13.16 | 1.52E-05 | 1.92E-03 |
| negative regulation of gene silencing by miRNA (GO:0060965) | 4 | 0.32 | 12.47 | 5.15E-04 | 2.94E-02 |
| response to cholesterol (GO:0070723) | 6 | 0.49 | 12.26 | 2.17E-05 | 2.47E-03 |
| negative regulation of posttranscriptional gene silencing (GO:0060149) | 4 | 0.34 | 11.85 | 6.10E-04 | 3.35E-02 |
| negative regulation of gene silencing by RNA (GO:0060967) | 4 | 0.34 | 11.85 | 6.10E-04 | 3.34E-02 |
| miRNA metabolic process (GO:0010586) | 4 | 0.37 | 10.77 | 8.36E-04 | 4.20E-02 |
| heterochromatin organization (GO:0070828) | 7 | 0.66 | 10.63 | 1.00E-05 | 1.39E-03 |
| response to sterol (GO:0036314) | 6 | 0.57 | 10.45 | 4.78E-05 | 4.74E-03 |
| production of miRNAs involved in gene silencing by miRNA (GO:0035196) | 5 | 0.49 | 10.21 | 2.30E-04 | 1.64E-02 |
| ventricular septum morphogenesis (GO:0060412) | 7 | 0.69 | 10.11 | 1.34E-05 | 1.73E-03 |

**Table 4:** Overrepresented Gene Ontology molecular functions ranked by fold enrichment value.

| GO molecular function complete | Number of Genes in List | Expected Number of Genes | Fold Enrichment | P Value | FDR |
| --- | --- | --- | --- | --- | --- |
| 5'-deoxyribose-5-phosphate lyase activity (GO:0051575) | 3 | 0.1 | 29.62 | 3.54E-04 | 3.25E-02 |
| transforming growth factor beta-activated receptor activity (GO:0005024) | 4 | 0.22 | 18.23 | 1.50E-04 | 1.63E-02 |
| activin binding (GO:0048185) | 4 | 0.25 | 15.8 | 2.38E-04 | 2.32E-02 |
| 1-phosphatidylinositol-3-kinase regulator activity (GO:0046935) | 4 | 0.27 | 14.81 | 2.93E-04 | 2.75E-02 |
| transforming growth factor beta binding (GO:0050431) | 5 | 0.39 | 12.88 | 8.80E-05 | 1.05E-02 |
| transmembrane receptor protein serine/threonine kinase activity (GO:0004675) | 4 | 0.32 | 12.47 | 5.15E-04 | 4.47E-02 |
| SMAD binding (GO:0046332) | 9 | 1.37 | 6.58 | 1.87E-05 | 3.73E-03 |
| catalytic activity, acting on DNA (GO:0140097) | 16 | 3.61 | 4.43 | 1.64E-06 | 6.52E-04 |
| protein phosphatase binding (GO:0019903) | 11 | 2.57 | 4.29 | 9.16E-05 | 1.07E-02 |
| phosphatase binding (GO:0019902) | 13 | 3.33 | 3.91 | 5.26E-05 | 6.80E-03 |
| kinase regulator activity (GO:0019207) | 13 | 3.82 | 3.41 | 1.93E-04 | 1.92E-02 |
| protein serine/threonine kinase activity (GO:0004674) | 21 | 7.43 | 2.83 | 3.28E-05 | 5.60E-03 |
| DNA-binding transcription activator activity, RNA polymerase II-specific (GO:0001228) | 21 | 7.46 | 2.81 | 3.49E-05 | 5.76E-03 |
| DNA-binding transcription activator activity (GO:0001216) | 21 | 7.53 | 2.79 | 3.96E-05 | 5.91E-03 |
| protein kinase activity (GO:0004672) | 25 | 9.89 | 2.53 | 3.54E-05 | 5.46E-03 |

### PPI Network Analysis

Following PANTHER analysis, STRING software was used to construct a network of the protein protein interactions (PPI) of the target genes. All 349 target genes were mapped by STRING against various protein databases. There were 783 interactions between nodes (genes) with an average node degree of 4.49. The expected number of intersections was only 543 so the network had significantly more interactions than would be expected (p < 1.0E-16). The network was exported to Cytoscape for further analysis via the Network Analyzer and MCODE tools.

**Figure 3:**
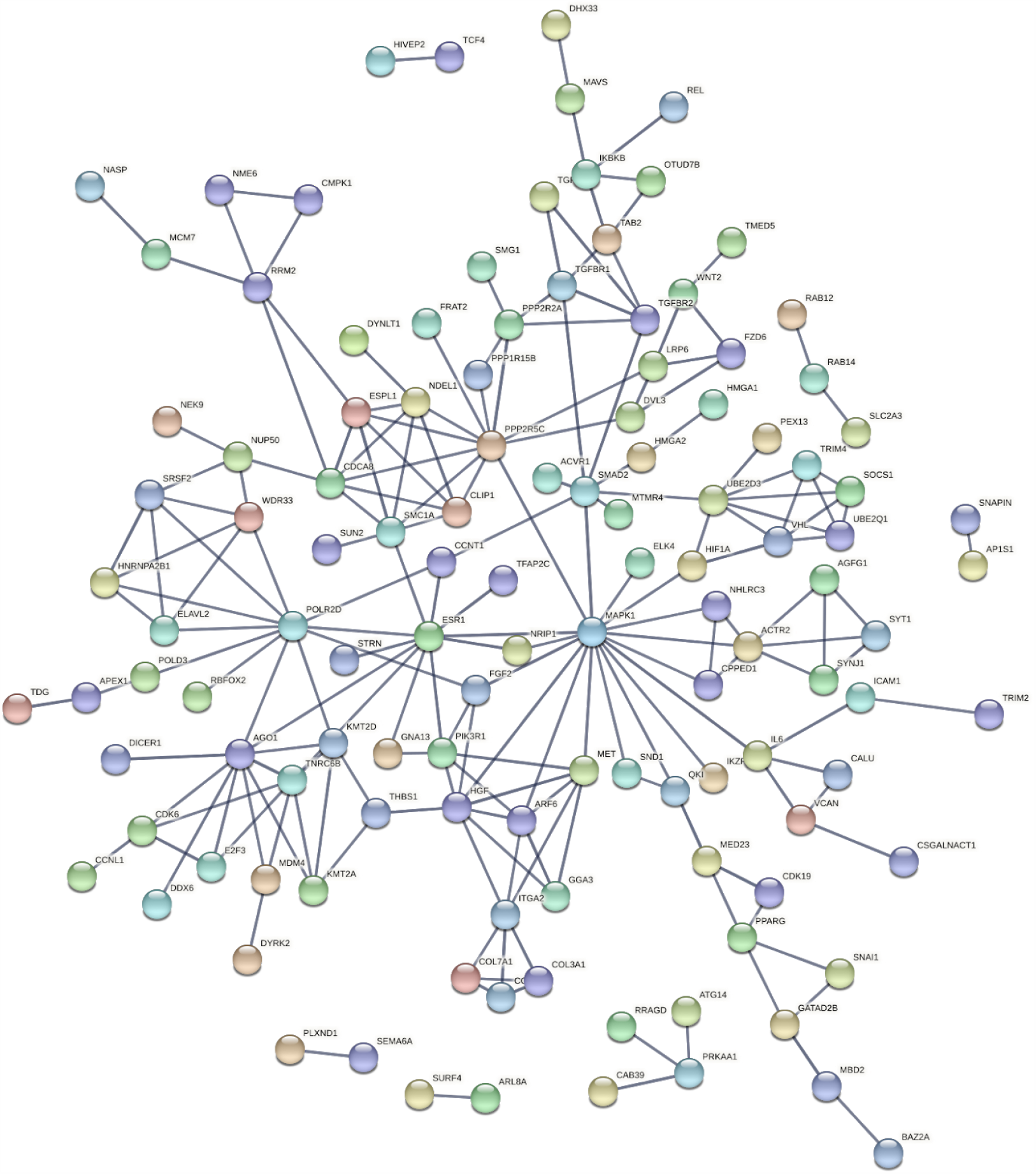
Protein protein interaction graph generated by STRING. Disconnected (isolated) nodes are hidden and minimum interaction confidence is set to 0.7 for clarity. Each circle represents one gene and its protein product. Lines between circles are “edges” and represent interactions.

**Figure 4:**
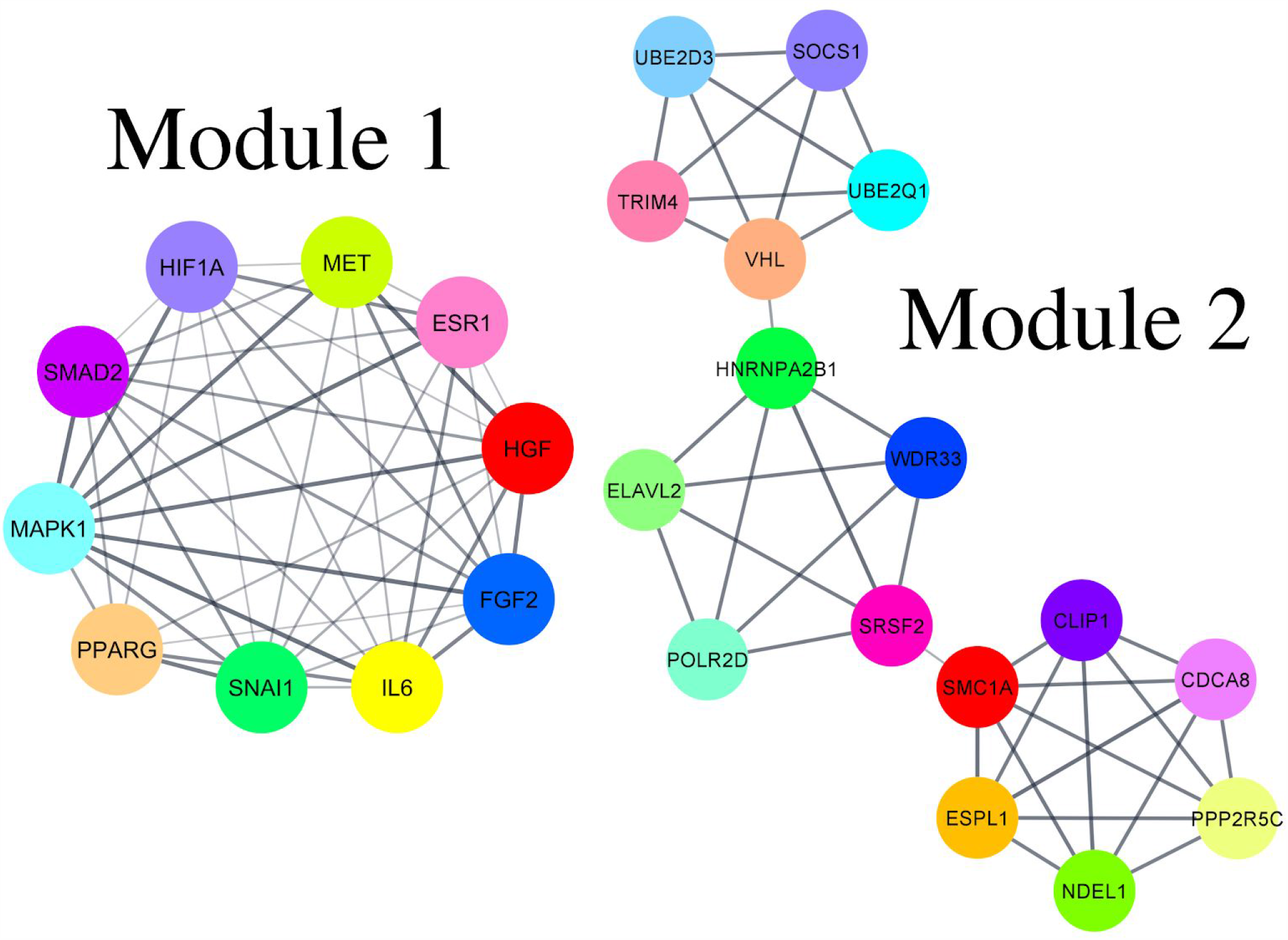
The two most significantly interconnected clusters of genes are displayed. Colors are arbitrary and each line between genes represents an interaction. Module 1 consists of 10 genes with 43 interactions and Module 2 consists of 16 genes with 37 interactions.

MCODE was used to detect modules (highly interconnected clusters of genes) in the Cytoscape network. 8 modules were identified by the MCODE algorithm. The top two modules by MCODE score are displayed. Module 1 was composed of 10 genes with 43 interactions between them. The genes were MAPK1, PPARG, SNAI1, IL6, FGF2, HGF, ESR1, MET, HIF1A, and SMAD2. Functional enrichment analysis of the module revealed that nine out of the ten genes (not SNAI1) were involved in the KEGG pathway “Pathways in cancer”. All ten genes were part of the Gene Ontology process “positive regulation of transcription, DNA-templated” and the Reactome Pathway of “signal transduction”. Module 2 was comprised of 16 genes with 37 interactions between them. Functional enrichment analysis showed that 6 of the genes were involved in chromatid separation, an important step of mitosis. Next, the tool Network Analyzer was used to select hub genes (minimum node degree 20). 11 genes passed the hub gene cutoff criteria: IL6, ESR1, MAPK1, FGF2, SMAD2, SNAI1, DICER1, CDK6, HGF, PPARG, and H1F1A. IL6, ESR1, MAPK1, FGF2, SMAD2, SNAI1, and HGF are all present in Module 1.

### Hub Gene Survival Analysis

Kaplan-Meier survival plots were created for each of the 11 hub genes, based on clinical data from 1144 lung adenocarcinoma and squamous cell carcinoma patients’ tissue samples. Patients were split by the median expression value of each gene into a “low” expression group and a “high” expression group. Except for MAPK1, H1F1A, and PPARG, the genes’ survival curves were statistically significantly different between low expression and high expression NSCLC patients (p<0.05).

**Figure 5:**
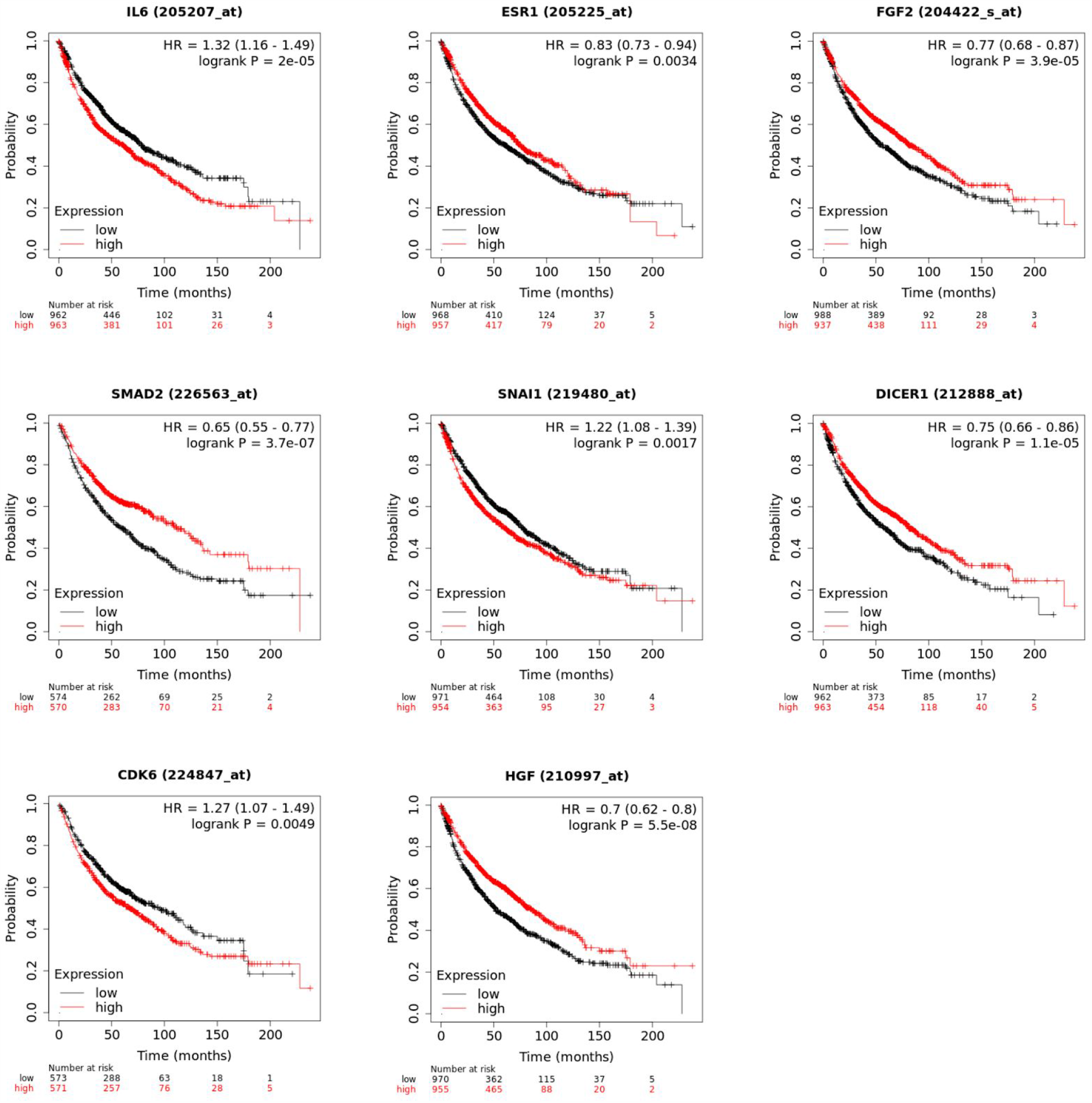
Kaplan Meier survival plots for IL6, ESR1, FGF2, SMAD2, SNAI1, DICER1, CDK6, and HGF. The black curve on each plot represents survival of patients with low expression of the considered gene and the red curve represents survival of those with high expression. The x axis is in months and the y axis is the probability of survival. The hazard ratio is a measure of probability of survival of the low expression group divided by the probability of survival of the high expression group. A hazard ratio of 1 indicates that the survival probability is the same for both groups.

Higher expression of IL6, SNAI1, and CDK6 were associated with poorer prognosis. Meanwhile, lower expression of ESR1, FGF2, SMAD2, DICER1, and HGF were linked to poorer prognosis. SMAD2 had the lowest hazard ratio (0.65) of all hub genes with a p value of 3.7E-07, indicating that differential expression of SMAD2 has a particularly large effect on patient survival. These 8 genes hold promise as prognostic biomarkers for NSCLC. Out of these nine genes, IL6 had the highest hub gene node degree from the protein protein interaction network.

**Figure 6:**
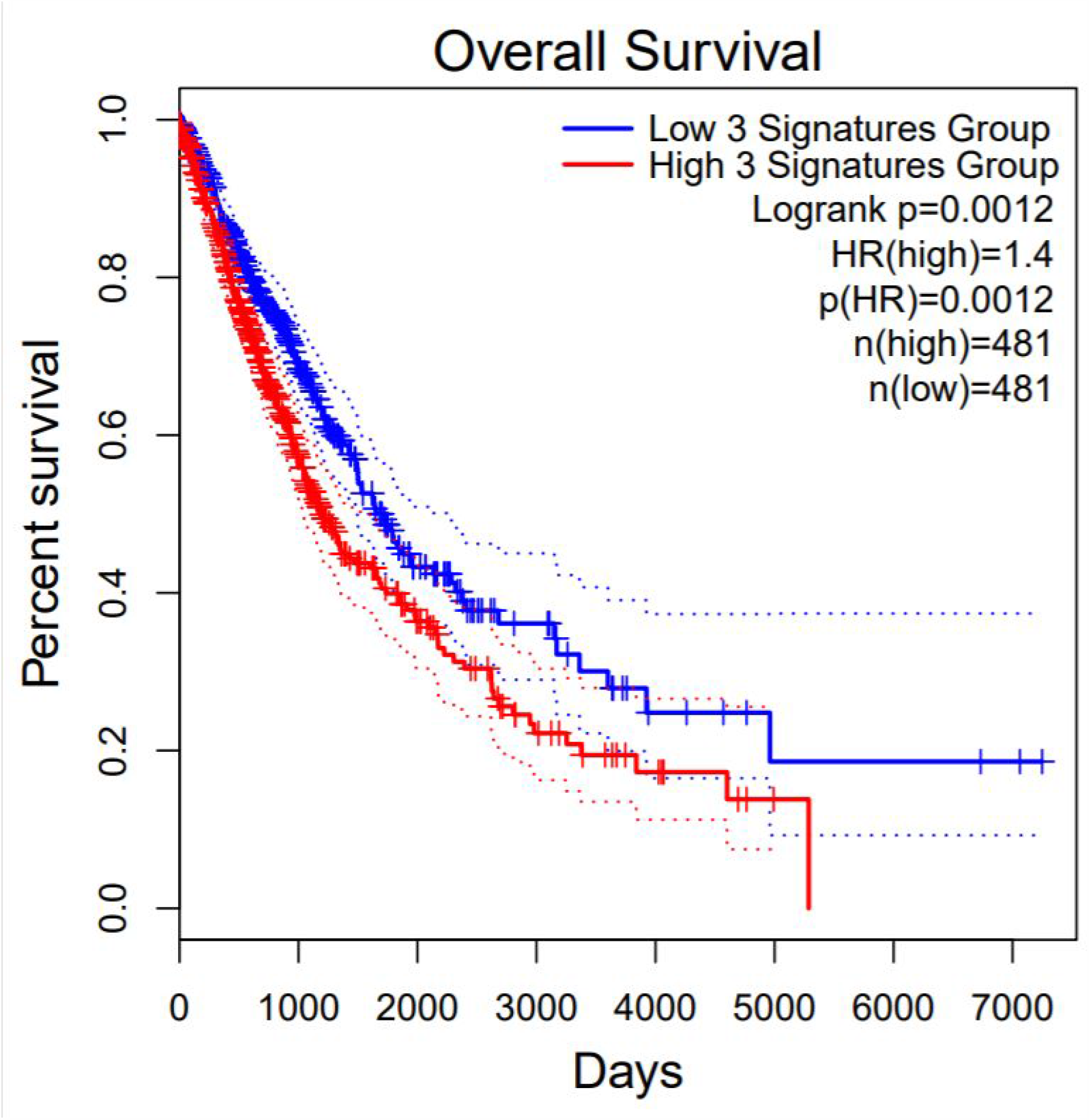
Survival plot from triple gene analysis using IL6 SNAI1, and CDK6. Blue indicated low expression while red indicates high expression. The x axis is time elapsed in days while the y axis is the survival percentage. Higher expression of these three genes was associated with poorer prognosis.

A triple gene survival analysis on NSCLC patients using a panel of IL6, SNAI1, and CDK6 was performed. The difference in high and low expression patients was statistically significant (p = 0.0012). The hazard ratio for the triple gene survival curve was higher than for each of the three genes alone, indicating that their combination lends prognostic power.

**Figure 7:**
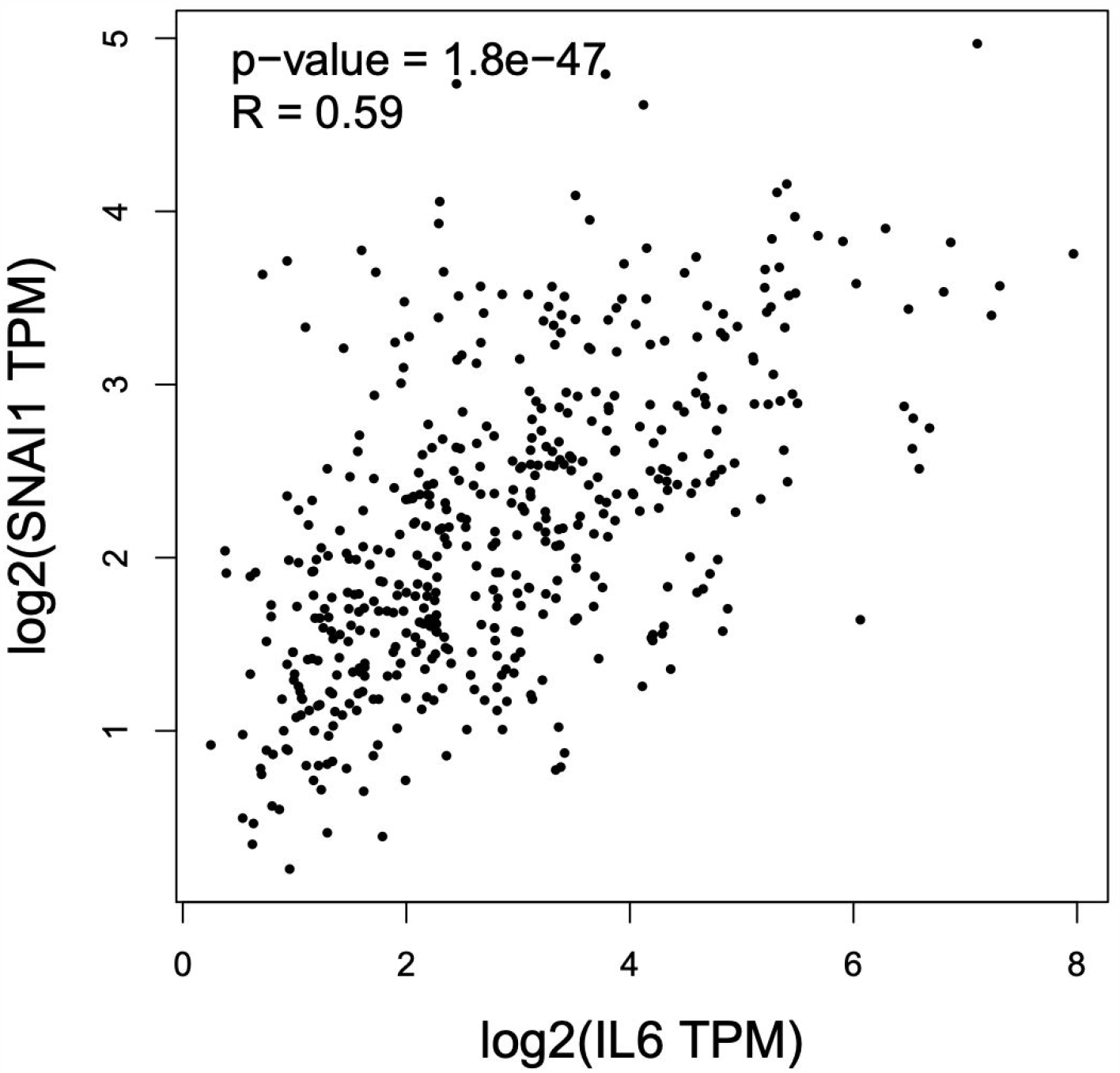
Scatter plot of IL6 and SNAI1 expression in LUAD patients with a log 2 transformation applied. A positive correlation is discernible. The p value and Spearman correlation values are displayed.

A correlation analysis of expression of IL6 and SNAI1 in TCGA LUAD patients showed significant correlation (p<0.01). The two genes were positively correlated with a Spearman coefficient of 0.59. GEPIA analysis demonstrated that IL6 and SNAI1 are underexpressed in the lungs of lung cancer patients based on data from TCGA patients.

### MiRNA Candidate Biomarker Selection

Kaplan-Meier survival analysis was performed for the 13 DEMiRNAs on 991 patients with either LUAD or LUSC. MiR-140, miR-29c, and miR-199a satisfied p < 0.05 and had hazard ratios of around 0.7, while the other ten miRNAs did not have significant hazard ratios. High expression of all three of these miRNAs was associated with better NSCLC outcome. Graphs of miRNA expression based on TCGA data in LUAD and LUSC patients were generated. MiR-140-3p was significantly underexpressed in tumor tissue in both LUAD and LUSC according to TCGA data analysis. MiR-29c-3p was significantly underexpressed in LUSC but was not significantly differentially expressed in LUAD (p>0.5). MiR-199a-5p was significantly overexpressed in both LUAD and LUSC. Receiver operating characteristic curves were generated for the three miRNAs. MiR-140-3p had an area under the ROC curve of 0.85 with p < 0.0001. MiR-199a-5p had an area of 0.6243 (p<0.0001) and miR-29c-3p had an area of 0.7443. Analysis of TCGA data showed differential methylation at MIR140 and MIR199A1, the genes that encode miR-140-3p and miR-199a-5p, respectively. Four cg probes near MIR140 were significantly hypermethylated in LUAD samples and seven cg probes near MIR199A1 were significantly hypermethylated in LUSC samples (p<0.05).

**Figure 8:**
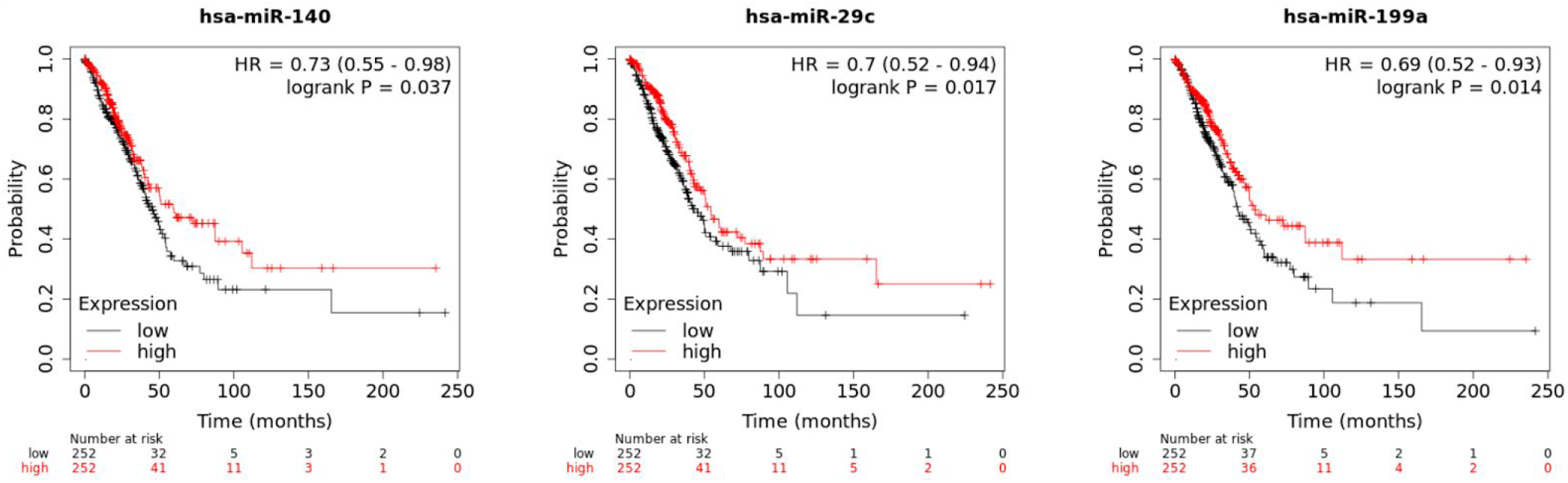
Kaplan Meier plots for miR-140, miR-29c, and miR-199a survival. The black curve represents patients with low expression in tissue of the considered miRNA while the red curve represents patients with high expression. The x axis is in months and the y axis is the probability of survival. The hazard ratio is a measure of probability of survival of the low expression group divided by the probability of survival of the high expression group. A hazard ratio of 1 indicates that the survival probability is the same for both groups.

**Figure 9:**
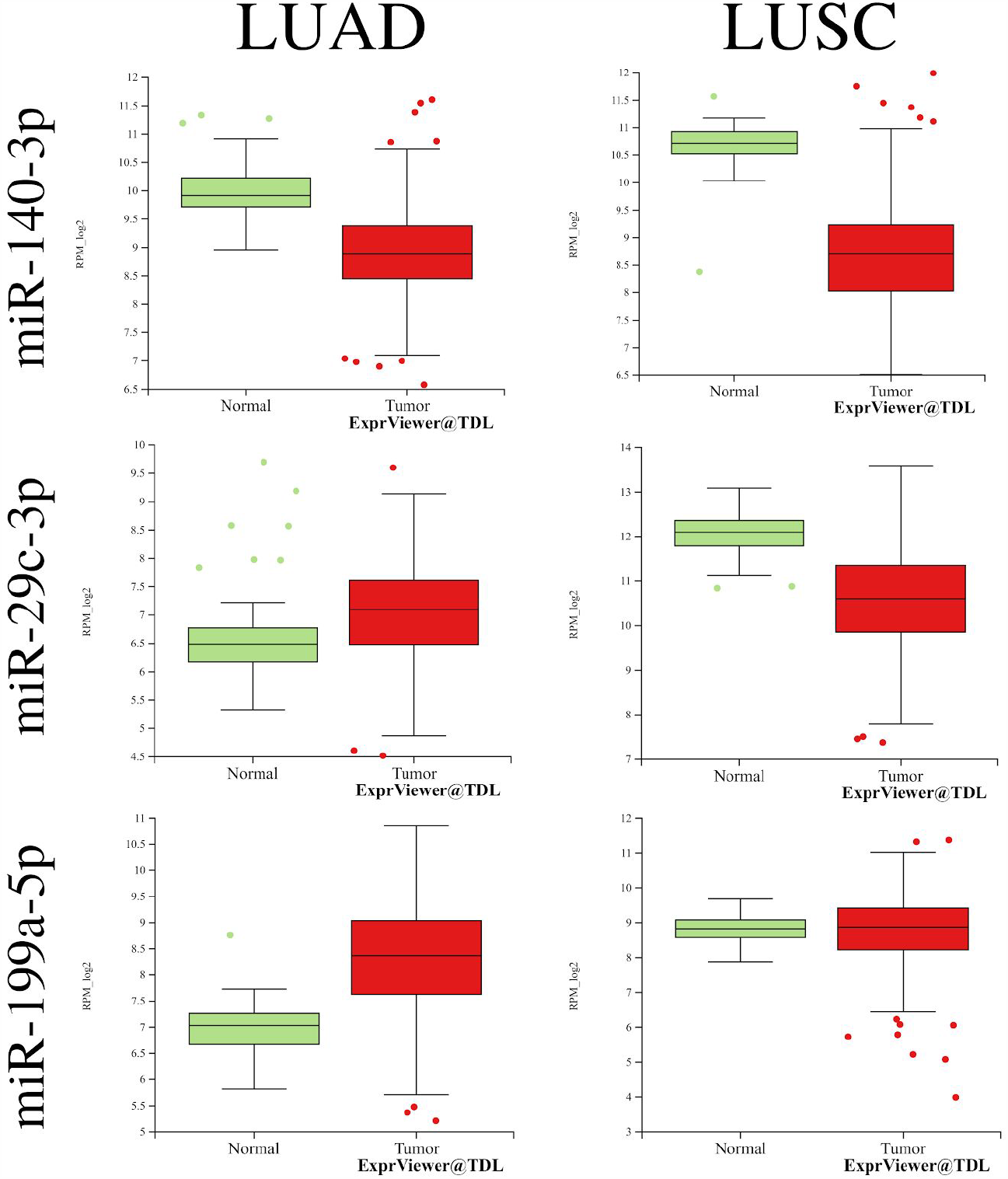
Expression of miR-140-3p, miR-29c-3p, and miR-199a-5p in LUAD and LUSC patients compared to controls. Results are shown in box plots with outliers marked separately as points.

**Figure 10:**
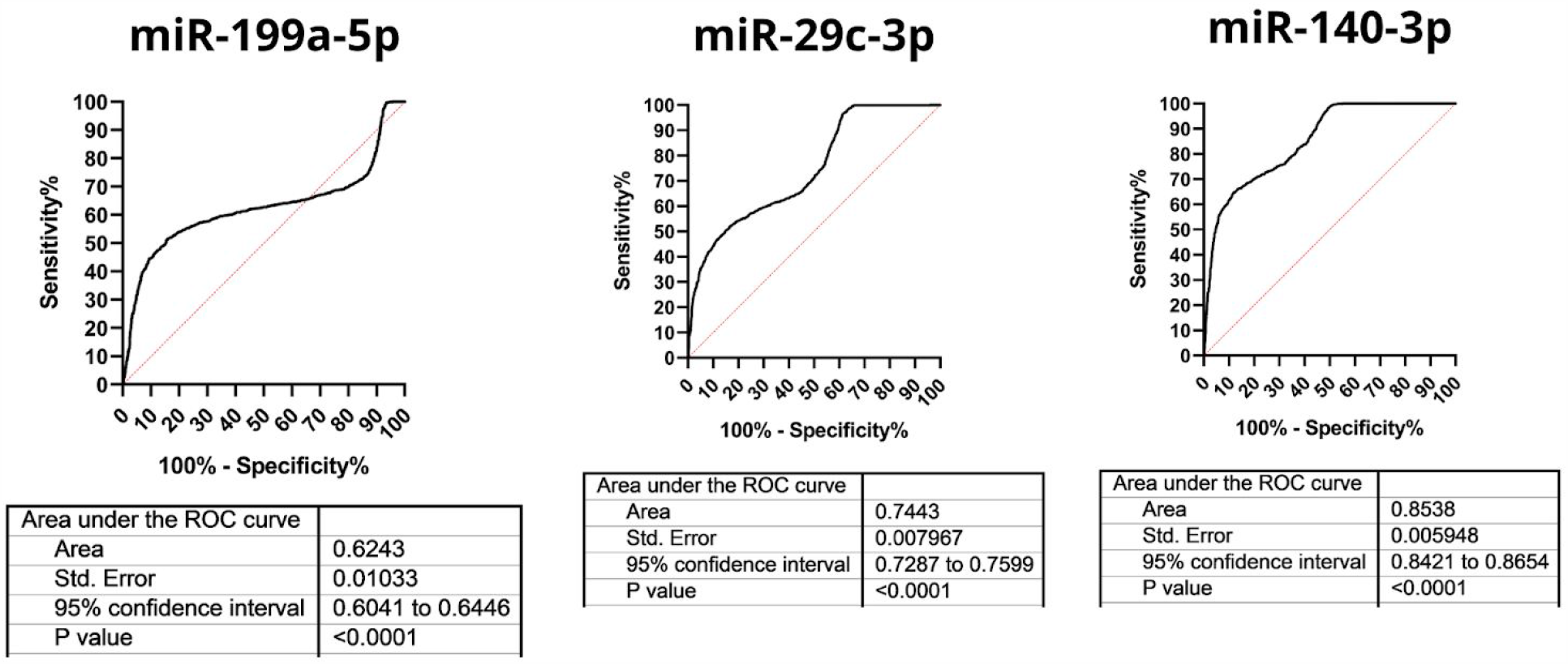
ROC graphs for miR-140-3p, miR-199a-5p, and miR-29c-3p based on expression data from 2178 controls and 1566 NSCLC patients.. ROC metrics are displayed in the tables. An area under the curve of 1 is ideal.

**Figure 11:**
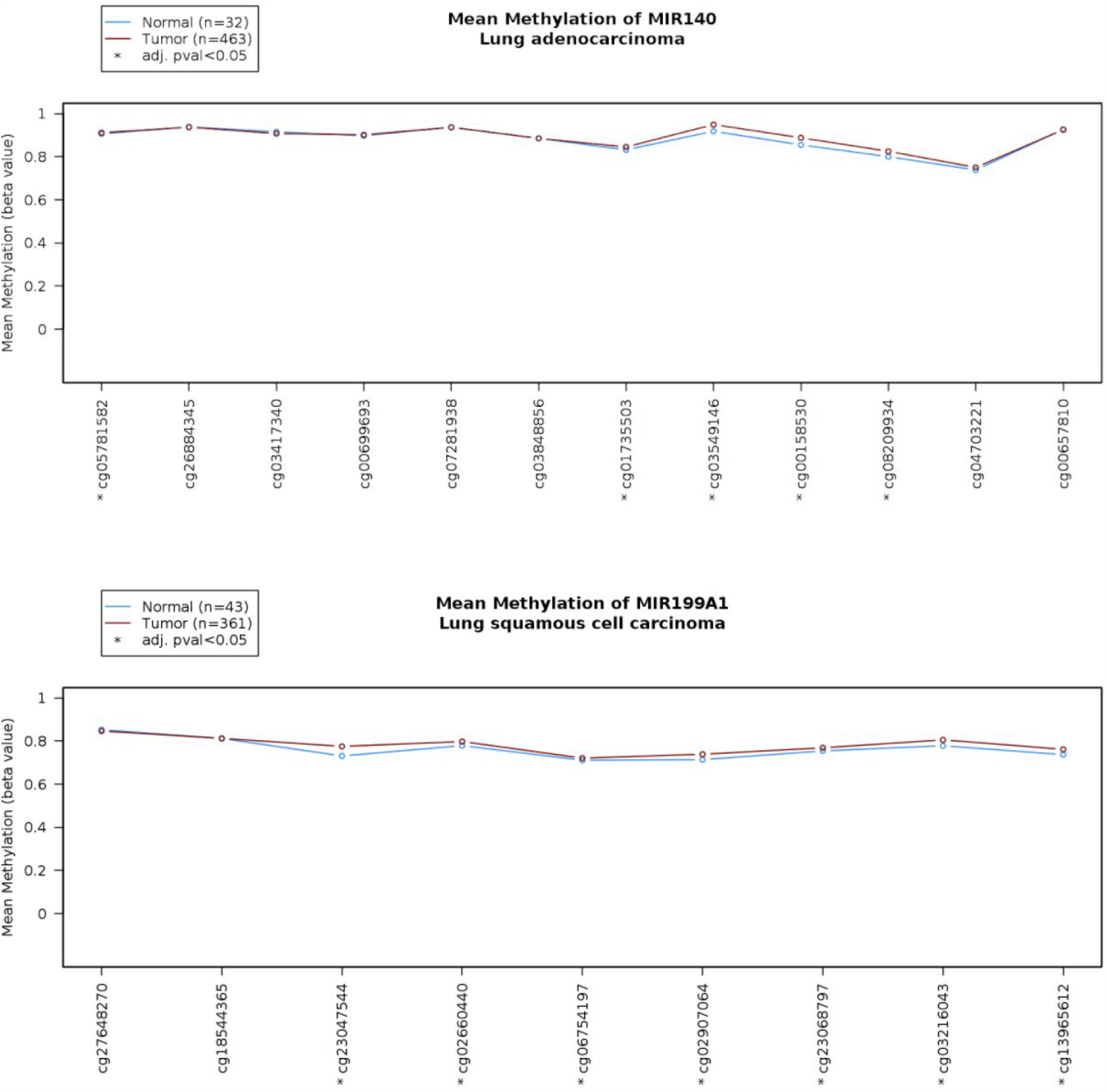
TCGA methylation analysis shows hypermethylation at cg probe sites near MIR140 and MIR199A1 in LUAD and LUSC respectively. The x axis consists of several nearby cg probes and the y axis represents the mean methylation beta value. A greater value indicates more methylation. Blue points represent control methylation levels while red points represent LUAD methylation levels.

## Discussion

Despite advances in non small cell lung cancer (NSCLC) treatments, the mortality rate has not significantly decreased. This is partly due to the lack of sensitive and accurate diagnostic tests, especially for early stage NSCLC. Analysis of dysregulated microRNAs and mRNAs in the blood and tissue of lung cancer patients may yield potential diagnostic and prognostic biomarkers, as well as improve understanding of the mechanisms of NSCLC. In the present study, four NSCLC miRNA expression datasets were screened and thirteen overlapping differentially expressed miRNAs were identified. 349 gene targets were predicted for the miRNAs. Network analysis and survival analysis were performed on the genes. Additionally, survival analysis was performed on the miRNAs.

Functional enrichment analysis of the 349 target genes revealed several overrepresented biological processes and molecular functions. The most highly enriched process was “positive regulation of epithelial to mesenchymal transition involved in endocardial cushion formation” (GO:1905007). The epithelial to mesenchymal transition process is a hallmark of cancer metastasis [33]. Next, the process of “pathway-restricted SMAD protein phosphorylation” was enriched by a factor of 18.23. SMAD proteins are signal transducers for transforming growth factor protein receptors which play important roles in cell cycle control, development, and growth. Impaired SMAD signaling has been linked to cancer and causes dysregulated cell growth [34-36]. SMAD3, a member of the SMAD family, was identified as a hub gene by network analysis and had the lowest hazard ratio (0.67) of all hub genes. The process of “heterochromatin assembly” was enriched by a factor of 13.16. Changes in heterochromatin can have large effects on transcription of certain genes and can be altered by cancer, contributing to genome instability [37-38]. Finally, the process of “negative regulation of gene silencing by miRNA” was enriched by 12.47 times. This may be significant as all thirteen overlapping differentially expressed miRNAs identified in this study were underexpressed, matching the process of “negative regulation of gene silencing by miRNA”.

Overrepresented molecular functions were related to protein kinase activity, transcription regulatory activity, chromatin binding, and transcription factor binding. The most highly enriched molecular function was GO:0051575: 5’-deoxyribose-5-phosphate lyase activity. These enzymes are indispensable in base excision repair, an important DNA repair mechanism, that is a line of defense against cancer development [39-40]. Analysis of Gene Ontology cellular components demonstrated that the RISC complex was enriched by a factor of 16.92. The RISC (RNA-induced silencing) complex uses miRNA to detect target mRNAs which it then cleaves [41]. RISC is essential for the function of miRNAs and its involvement in NSCLC demonstrates the influential role of epigenetic control of gene expression. Overall, these Gene Ontology processes and functions elucidate the biological mechanisms that the thirteen underexpressed DEMiRNAs influence and shed light on the importance of SMAD signaling and other mechanisms in NSCLC.

IL6, ESR1, MAPK1, FGF2, SMAD2, SNAI1, DICER1, CDK6, HGF, and H1F1A were identified as hub genes by Cytoscape analysis due to having a node degree of greater than 20. Several of these genes are well known to be implicated in cancer, namely IL6 and MAPK1. Kaplan Meier survival analysis of these eleven genes showed that all but MAPK1 and PPARG had significant hazard ratios. These nine significant genes are proposed as blood-based mRNA biomarkers for non-small cell lung cancer. Their presence in blood should be experimentally validated.

Meanwhile, miR-140, miR-29c, and miR-199a were selected as promising miRNA candidate biomarkers following Kaplain Meier analysis.

MiR-140-3p has been demonstrated to play a role in cancer and is considered a tumor suppressor. This is supported by the results of this present study which demonstrated that miR-140-3p was underexpressed in four separate NSCLC datasets. Tumor suppressors are generally underexpressed in cancer to allow tumors to develop. Huang et al. discovered that miR-140-3p was underexpressed in the tissue of SCLC patients and was correlated with survival and tumor stage [42]. Additionally, the presence of miR-140-3p was found to inhibit cell proliferation of SCLC cells and invasion. They demonstrated that the BRD9 gene is directly regulated by miR-140-3p and promotes SCLC proliferation. BRD9, or Bromodomain-Containing Protein 9, is involved in chromatin remodeling and transcriptional regulation, two processes identified as overrepresented by the present study’s functional enrichment analysis. Patients with greater levels of BRD9 had worse prognosis. Thus, the BRD9/miR-140-3p regulatory axis may be a potential therapeutic target in SCLC treatment. Another study, by Kong et al. demonstrated that mir-140-3p “inhibits proliferation, migration, and invasion of lung cancer cells by targeting ATP6AP2” [43]. In their study, miR-140-3p was underexpressed in lung cancer tissue compared to normal lung tissue. They determined that miR-149-3p directly downregulates the expression of ATP6AP2 by binding to the 3’ UTR. The authors propose that dysregulation of the miR-140-3p/ATP6AP2 axis may disrupt the Wnt signaling pathway and promote lung cancer. Another ATP-related protein, ATP8A1, was found to be downregulated by miR-140-3p in NSCLC. Dong et al. reported that the addition of miR-140-3p to NSCLC cells inhibited growth and decreased expression of ATP8A1 [44]. MiR-140-3p has been implicated to have a tumor suppressor role in several other cancers by many studies. MiR-140 “promotes cancer stem cell formation” in breast cancer when underexpressed, induces apoptosis in colorectal cancer cells, suppresses cervical cancer metastasis, and inhibits the proliferation of bladder cancer cells. TCGA analysis revealed that the MIR140 gene, which encodes miR-140-3p, is hypermethylated at four nearby CpG sites in LUAD tissue, indicating decreased expression due to methylation. This is consistent with the findings that miR-140-3p is underexpressed in NSCLC based on differential expression analysis. MiR-140-3p shows promise as a therapeutic target and diagnostic/prognostic biomarker. It is underexpressed in NSCLC patients in all four datasets examined as well as TCGA data and has a hazard ratio of 0.73 (p<0.05). Additionally, the MIR140 gene is significantly hypermethylated. ROC analysis on miRNA expression data from GSE137140 showed that miR-140-3p has an AUC of 0.85 which is highly promising.

MiR-29c-3p was also selected as a candidate biomarker. Chen et al. found that miR-29c-3p inhibits colon cancer cell invasion and Wu et al. found that it suppresses hepatocellular carcinoma tumor progression [45-46]. Fang et al. reported that underexpression of miR-29c-3p is “associated with a poor prognosis” in laryngeal squamous cell carcinoma [47]. Additionally, Van Sinderen et al. showed that restoring levels of miR-29c-3p in endometrial cancer cells led to reduced growth [48].

MiR-199a-5p is also implicated in cancer. Chen et al. describe the tumor-suppressing effects of miR-199a-5p in triple negative breast cancer [49]. Additionally, Li et al. report that this miRNA “suppresses non-small cell lung cancer via targeting MAP3K11” and Zhu et al. found that it can inhibit the growth of colorectal cancer cells [50-51]. Interestingly, Ma et al. demonstrated that “miR-199a-5p inhibits the progression of papillary thyroid carcinoma by targeting SNAI1” [52]. SNAI1 is one of the eight hub genes with statistically significant prognostic value. TCGA analysis showed that MIR199A1, the gene that encodes miR-199a-5p, is hypermethylated in LUSC, indicating decreased expression.

MiR-140-3p, miR-29c-3p, and miR-199a-5p are all well-documented to play protective roles in several types of cancer, including NSCLC. They are reported to have tumor suppressive functions and are usually downregulated in cancer. Existing literature has described the potential therapeutic value of these miRNAs, however, to the author’s knowledge, these miRNAs have not been considered for use in blood-based diagnostic and prognostic tests. The main contribution of the present study is that miR-140-3p, miR-29c-3p, and miR-199a-5p are potential non-invasive biomarkers for NSCLC.

One limitation of this study was that no normalization was performed between datasets (although this is common with gene expression meta-analyses). Also, this analysis did not distinguish between males and females. There may be relevant differences in gene expression in NSCLC between sexes [53]. Another important caveat is that the research only compared healthy normal controls and NSCLC patients. However, NSCLC miRNA expression was not compared with that of other cancers or lung diseases, such as chronic obstructive pulmonary disorder. The miRNAs and mRNAs identified in this study as biomarkers are not necessarily able to discriminate between NSCLC and other diseases. A follow-up study comparing miRNA expression between NSCLC and other cancers and lung diseases is warranted.

An advantage of this study was that four studies were analyzed for differential miRNA expression, comprising 1978 NSCLC and 1932 control samples total. This large sample size accounts for NSCLC heterogeneity more adequately than an individual study can. Another advantage is that stringent miRNA target prediction standards were applied: three prediction tools were used to calculate target genes and only genes confirmed by all three were kept. Additionally, the differentially expressed miRNAs identified in this study are found in both plasma and serum according to GEO datasets, allowing for greater flexibility in screening.

Future research directions include further testing of patient blood samples to evaluate the diagnostic and prognostic power of the proposed miRNAs and mRNAs. The mRNAs should be validated to exist in circulating blood. Additionally, miR-140-3p, miR-29c-3p, and miR-199a-5p should be considered as viable therapeutic targets for NSCLC. Delivering supplemental doses of these miRNAs may have beneficial effects, as demonstrated in cell cultures.

## Conclusion

The present study identified 13 microRNAs that are underexpressed in the tissue and blood of non small cell lung cancer patients. MiR-140-3p, miR-29c, and miR-199a are candidate biomarkers based on bioinformatics analysis and demonstrated prognostic power. An ROC analysis of miR-140-3p expression between NSCLC patients and controls had an area under curve (AUC) value of 0.85, indicating significant discriminatory ability. Functional enrichment analysis of the miRNA target genes revealed several overrepresented pathways relevant to cancer, including SMAD signaling. Eight target genes were hub genes in the protein protein interaction network and possessed significant prognostic value. A combination of IL6, SNAI1, and CDK6 achieved a hazard ratio of 1.4 with p < 0.001.

The biomarkers proposed in the present study are especially valuable because not only are they differentially expressed in blood, but also in tissue. Since all analyzed miRNAs were underexpressed in both tissue and blood, detecting expression of a biomarker miRNA in blood may provide information on its expression in tumor tissue as well. Clinical validation of these computationally identified candidate miRNAs and mRNAs is warranted.

## Supporting information

Predicted Target Genes of 13 DEMiRNAs

Compiled LogFC Values for 13 DEMiRNAs

## Data Availability

Data is freely and publicly available from the Gene Expression Omnibus.

https://www.ncbi.nlm.nih.gov/geo/query/acc.cgi?acc=GSE137140

https://www.ncbi.nlm.nih.gov/geo/query/acc.cgi?acc=GSE94536

https://www.ncbi.nlm.nih.gov/geo/query/acc.cgi?acc=GSE53882

https://www.ncbi.nlm.nih.gov/geo/query/acc.cgi?acc=GSE93300

## Abbreviations

NSCLC: Non-small cell lung cancer
LUAD: Lung adenocarcinoma
LUSC: Squamous cell lung cancer
miRNA: microRNA
DEMiRNA: Differentially expressed microRNA

## Acknowledgements

My sincere thanks to Professor Eliezer Van Allen and the Van Allen lab for their generous help.

